# Limits and opportunities of SARS-CoV-2 antigen rapid tests – an experience based perspective

**DOI:** 10.1101/2020.09.22.20199372

**Authors:** Verena Schildgen, Sabrina Demuth, Jessica Lüsebrink, Oliver Schildgen

## Abstract

**Background:** Due to the steadily rising case numbers of SARS-CoV-2 infections worldwide there is an increasing need for reliable rapid diagnostic devices in addition to existing gold standard PCR-methods. Actually, public attention is focused on antigen assays including lateral flow tests (LFTs) as diagnostic alternative. Therefore, different LFTs were analyzed regarding their performance in a clinical setting.

**Material and Methods:** A pilot sample panel of 13 BALFs and 60 throat washing samples (TWs) with confirmed PCR results as well as 8 throat washes invalid by PCR was tested with the BIOCREDIT test (RapiGEN), the Panbio™ assay (Abbott), and the SARS-CoV-2 Rapid Antigen Test (Roche).

**Conclusion:** The analyzed antigen test showed an inter-assay correlation of 27.4% with overall specificities ranging from 19.4% to 87.1%, while sensitivities of the respective tests ranged between 33.3% and 88.1%. Although these assays did not entirely meet all high expectations their benefit has to be carefully evaluated for the respective test strategy and setting.

## Introduction

In December 2019 the public became aware of the new betacoronavirus SARS-CoV-2 due to an outbreak in Wuhan, China (1). Very soon it turned out that spreading of the virus cannot be prevented and Covid-19 was declared a pandemic in March 2020 (https://www.euro.who.int/en/health-topics/health-emergencies/coronavirus-covid-19/news/news/2020/3/who-announces-covid-19-outbreak-a-pandemic). In order to minimize the risk of infection different undirected as well as targeted tracking strategies were developed, whose success is dependent on extensive testing of the largest possible number of people (2). For SARS-CoV-2 detection different PCRs are used in routine diagnostic. Although these PCRs actually represent the diagnostic gold standard (3), valuable time passes until the result is available (https://www.nytimes.com/2020/08/04/us/virus-testing-delays.html). Since it was shown that rapid antibody screenings are not suitable to evaluate chains of infection or their interruption, other kinds of rapid on-site tests are needed to perform the requested mass testing.

Among these are the BIOCREDIT COVID-19 Ag test of RapiGEN (Korea), the Panbio™ COVID-19 Ag of Abbott (Germany), and the SARS-CoV-2 Rapid Antigen Test of Roche (Germany). These lateral flow tests (LFTs) are based on immunochromatography and show SARS-CoV-2 antigen presence by a coloured test line. Sampling should be performed in the nasopharynx with a supplied swab, but except the Panbio™ assay UTM/VTM (universal/viral transport media) are also appropriate. Moreover it has been shown that specimen other than nasopharyngeal swaps are superior regarding diagnostic sensitivity (4, 5). For this reason we tested a pilot sample panel of 60 throat washes and 13 bronchoalveolaer fluids (BALFs) regarding its test performance.

## Material and Methods

A pilot sample panel of 13 BALFs and 60 throat washing samples (TWs) with confirmed PCR results (RealStar® SARS-CoV-2 RT-PCR Kit, Altona, Germany) as well as 8 throat washes invalid by PCR was tested with three SARS-CoV-2 antigen lateral flow tests (LFTs). We compared the BIOCREDIT assay of RapiGEN, the Panbio™ assay of Abbott, and the SARS-CoV-2 Rapid Antigen Test of Roche. If supplied, controls were performed according to the manufacturer. For test procedure sample volumes of 55µl (Roche), 110 µl (Abbott), and 150 µl (RapiGen) BALF/TW instead of extracted swap material were applied to the respective assays, but otherwise tests were performed according to the manufacturers’ recommendations. Additional analysis with the above listed tests were performed with PCR invalid samples (n=8).

## Results

First, proper performance of the Panbio™ was confirmed with the supplied controls (fig. 1A). After pooling these controls were applied to the devices of Roche, which also detected the positive control, and of RapiGEN, which was negative for the Abbott controls (fig.1B). In order to figure out if SARS-CoV-2 antigen can be detected in specimens other than nasopharyngeal swaps we applied either diluted or original PCR positive specimen on the device and checked for the presence of the control line (C) and the test line (T). To increase the probability of antigen detection samples with Ct values <16 for E- and S-gene were used. Although there was not sufficient material of sample II, this pilot approach showed that the specimens used allow a proper test performance and that SARS-CoV-2 antigen can principally be detected in throat washes (I and II) and BALF (III) (fig. 2).

**Figure 1:**
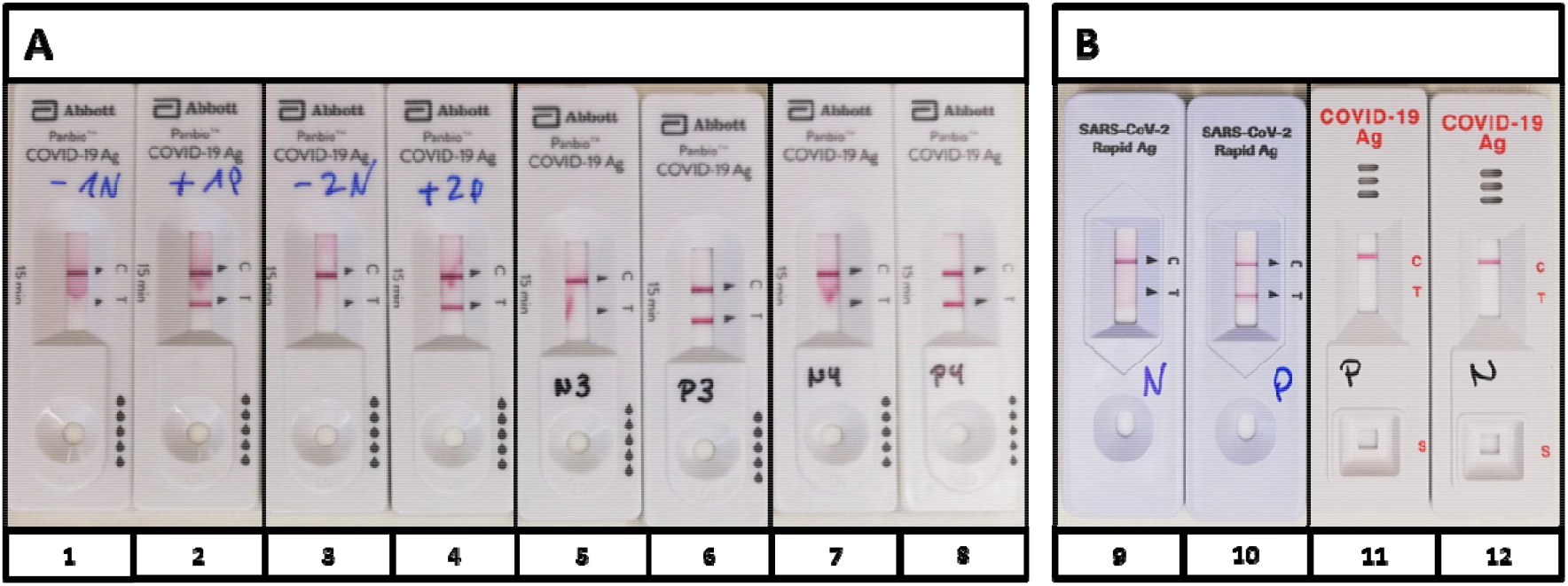
Controls of the Panbio™ COVID-19 Ag test (Abott). **A** Supplied PCs (2, 4, 6, 8) and NCs (1, 3, 5, 7) of four kits were performed according to the manufacturer’s protocol. **B** Controls of approach A were pooled and applied to the antigen tests of Roche (9, 10) and RapiGEN (11, 12). While the PC of Abbott also became positive with Roche (10), the BIOCREDIT test did not recognize the positive control (11). PC = positive control, NC = negative control

**Figure 2:**
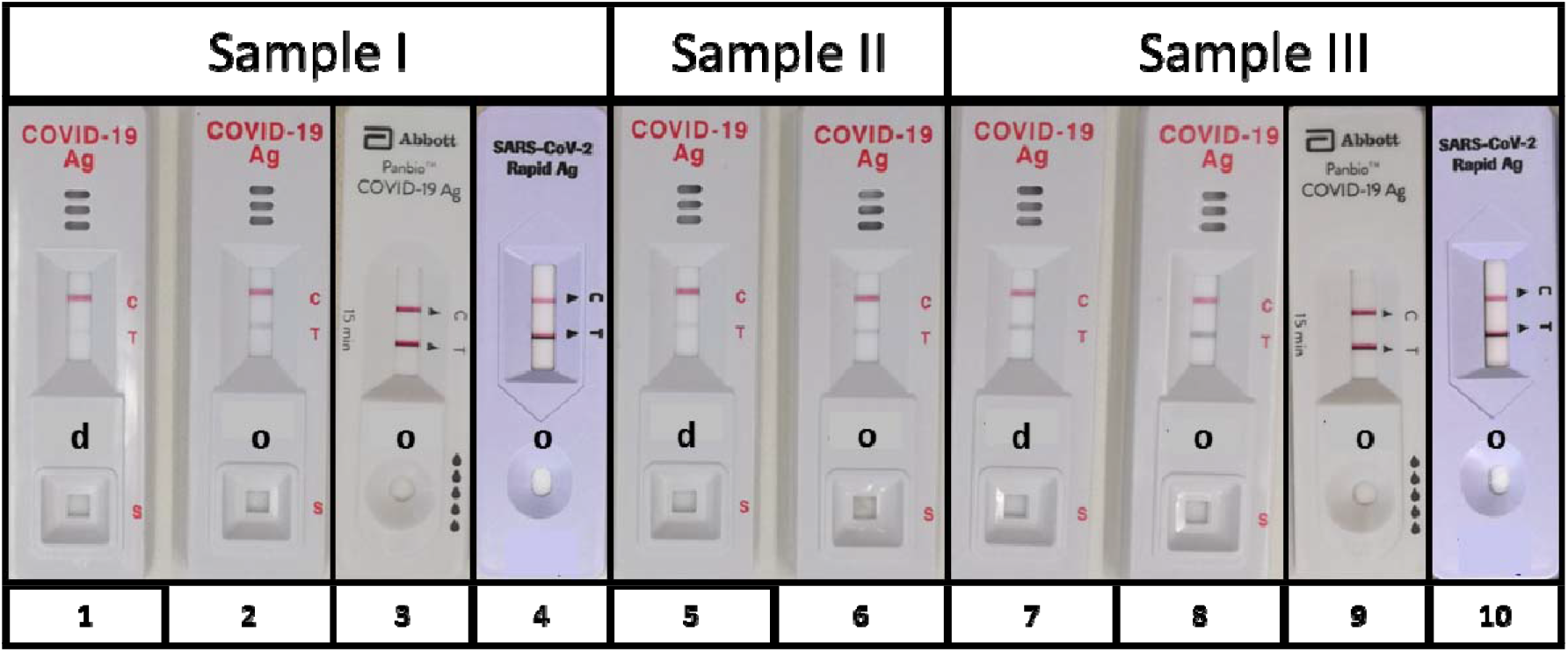
SARS-CoV-2 antigen detection. To evaluate the suitability of specimen others than nasopharyngeal swaps two PCR positive throat washes (I: Ct_E-Gen_=15,6, Ct_S-Gen_=14,8; II: CT_E-Gen_=14,7, Ct_S-Gen_=14,9) and one BALF (III: Ct_E-gen_=13,1, Ct_S-Gen_=12,6) were used for initial evaluation of lateral flow test performance. SARS-CoV-2 antigen was detected with the BIOCREDIT test in all samples, but dilution in assay buffer (d: 1, 5. 7) decreases sensitivity compared to original fluid (o: 2, 6, 8). Undiluted samples I and III were also used for an initial analysis with the Panbio™ COVID-19 Ag test (Abott) (3, 9) and the SARS-CoV-2 Rapid Antigen Test (Roche) (4, 10) also delivering positive results in both tests. As shown by the control line the specimens allowed a proper test performance and did not contain any inhibitory substances. d=1:2 diluted with supplied buffer, o= original

The analysis of the complete test cohort revealed overall sensitivities of 33.3% (RapiGEN), 50% (Abbott), and 88.1% (Roche) with opposite overall specificities of 87.1% (RapiGEN), 77.4% (Abbott), and 19.4% (Roche) (table 1). This means positive predictive values (PPVs) ranging from 59.7% (Roche) to 77.8% (RapiGEN) and negative predictive values (NPVs) of 53.3% (Abbott), 54.6% (Roche) and 49.1% (RapiGEN).

**Table 1:** Sensitivity* and specificity* of SARS-CoV-2 antigen detection in PCR tested specimen. Three LFTs were used for evaluation. Analysis of the complete test cohort (n=73) reveals overall sensitivities of 33.3% (RapiGEN), 50.0% (Abbott), and 88.1% (Roche). Overall specificities range from 19.4% (Roche) to 77.42% (Abbott) up to 87.1% (RapiGEN). 50 samples confirmed symptomatic or asymptomatic reveal highest sensitivities (84.6% and 100%), but poor specificities with Roche, whereas highest specificities (76.9% and 92.9%) are reached by RapiGEN with sensitivities of 30% respectively. In total the three LFTs match in 20 of 73 samples (27.4%). * in percent, BAL= bronchoalveolar lavage, TW= throat wash

|  | RapiGEN |  | Abbott |  | Roche |  |
| --- | --- | --- | --- | --- | --- | --- |
|  | Sensitivity | Specificity | Sensitivity | Specificity | Sensitivity | Specificity |
| BAL<br>N=13 | 54.6 | 100 | 72.7 | 100 | 81.82 | 100 |
| TW<sub>symptomatic</sub><br>N=23 | 30.0 | 76.9 | 40.0 | 84.6 | 100 | 7.7 |
| TW<sub>asymptomatic</sub><br>N=27 | 30.8 | 92.9 | 38.5 | 71.4 | 84.6 | 14.3 |
| Total<br>N=73 | 33.3 | 87.1 | 50.0 | 77.4 | 88.1 | 19.4 |

The analysis of 50 samples confirmed symptomatic (46%) or asymptomatic (54%) with regard to the respective PCR result, led to sensitivity/specificity values in asymptomatic individuals of 30.8/92.9% (RapiGEN), 38.5/71.4% (Abbott), and 84.6/14.3% (Roche) and to sensitivity/specificity values in symptomatic individuals of 30.0/76.9% (RapiGEN), 40.0/84.6% (Abbott), and 100/7.7% (Roche). Regarding the BALF specimens the assays of Abbott and Roche showed performance data of 72.7% or 81.8% sensitivity and 100% specificity, whereas RapiGEN also showed 100% specificity but 54.6% sensitivity (table 1).

When comparing the antigen tests with regard to their correlation it turned out that the RapiGEN-Roche coincidence was 35.6% (26 samples), the RapiGEN-Abbott coincidence 64.4% (47 samples), and the Roche-Abbott coincidence 50.7% (37 samples). In total, the LFTs provided the same results in 20 out of 73 samples (27.4%). These results damped the expectation that PCR invalid samples could reliably be analyzed with any of these rapid antigen tests, especially as two of these samples (n=8) still remained invalid in one LFT, respectively, and only three samples coincide as negative in all LFTs.

When checking any correlation of viral RNA load and presence of SARS-CoV-2 antigen we were able to monitor two asymptomatic individuals, who were PCR positive for more than five weeks before recovering in week six. Ct values ranged from 27.9 to 34.3 in patient A (data series dark grey) and from 23.8 to 35.2 in patient B (data series light grey). The BIOCREDIT assay detected SARS-CoV-2 antigen only in three samples of patient A with Ct values >30 (figure 3). Sample four of patient B was only detected by the Panbio™ assay and sample 5 only by the SARS-CoV-2 Rapid Antigen Test of Roche. Sample 1 of patient A remained antigen negative in all tests, despite a positive PCR result, whereas none of the samples was identified as positive in all three assays.

**Figure 3:**
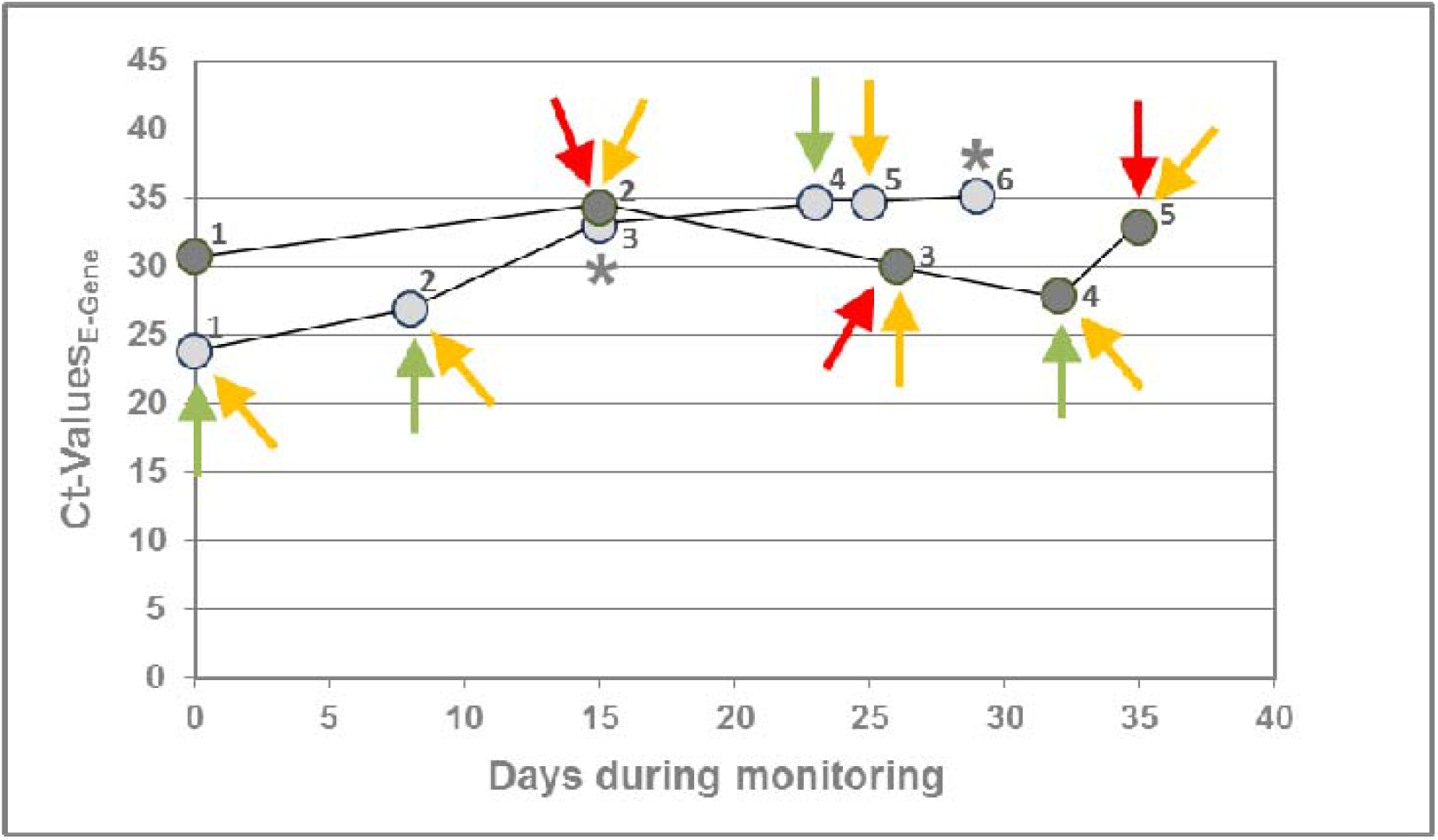
Correlation of SARS-CoV-2 antigen detection with RNA load. This figure shows longitudinal SARS-CoV-2 detection by PCR in two asymptomatic individuals (A: dark grey; B: light grey). In both cases SARS-CoV-2 RNA could be detected for about five weeks before recovering in week six. The BIOCREDIT assay (red) detected SARS-CoV-2 antigen only in patient A in three samples with Ct >30. Although the assays of Abbott (green) and Roche (yellow) identified 4 and 7 samples as positive, respectively, one sample remained antigen negative (sample 1 of patient A). Except the negative sample there was no coincidence between the LFTs at all. * only tested negative by BIOCREDIT due to insufficient material

## Discussion

Due to the pandemic spread of SARS-CoV-2 it would be desirable to possess reliable rapid tests for infection control by early notification of cases to enable an effective outbreak management. Therefore, R&D efforts are focused on rapid diagnostic tests including SARS-CoV-2 antigen tests.

Here we tested SARS-CoV-2 antigen assays by RapiGen, Abbott, and Roche. According to the manufacturers’ instructions, sampling should be performed with supplied swaps, but this test procedure limits test comparability as every swap represents an individual sample even if taken from the same patient. For this reason, and because UTM/VTM is also appropriate except for the Panbio™ assay, we evaluated assay performance with BALF and throat washes as these were performed with a physiological solution of NaCl (0.9% w/v) and thus did not contain any additional interfering chemicals compared to the allowed transport media. Although the Panbio™ assay excludes specimens other than swaps, it is unlikely that the Panbio™ buffer components ProClin300 and sodium acid (preservatives) or Tricin (buffer substance capturing divalent metal ions) have direct influence on SARS-CoV-2 antigen binding capacity. Solely missing Tween20 might have influenced antigen binding in the PanBio™ assay, but although test control lines occurred properly in all PCR pre-tested samples the overall correlation between all LFTs was only 27.4%.

The fact that the positive control supplied by Abbott was detected by Roche, but not by RapiGen, suggests that the assays may detect different antigens, which additionally complicates the estimation of test comparability and usability in clinical routine settings. Especially, as it still remains unclear if further common phenomena such as defective interfering particles, antigen drift or antigen shift occur during the current pandemic influence assay performance of any SARS-CoV-2 antigen test.

As SARS-CoV-2 antigen is detected in samples of individuals ranging from asymptomatic + PCR negative, to symptomatic + PCR negative, to asymptomatic + PCR positive, to symptomatic + PCR positive, antigen tests are not suitable for routine diagnostics as long the complex relationships between viral RNA load, SARS-CoV-2 antigen detection, and clinical symptoms remain unsolved. This conclusion is supported by the manufacturer’s recommendations, who explicitly claim that their assays are not approved as stand-alone diagnostic and the limitations that the assay should be performed “in patients with clinical symptoms” (RapiGEN) or that the test “is not intended to detect from defective (non-infectious) virus during the later stages of viral shedding that might be detected by PCR molecular tests” (Abbott). This means that these assays are not appropriate for the screening of asymptomatic individuals and cannot be recommended to date to be broadly used in any setting in which reliable diagnostics are crucial to avoid spreading of the virus, such as hospitals and long-term care facilities for the elderly or other risk groups, especially, as limited information on host and viral factors influencing shedding of SARS-CoV-2 antigens and their correlation to infectious viruses impede any prognosis on infectivity.

Although it actually seems that exclusive antigen tests have research character rather than being true IVDs and unfortunately cannot replace PCR assays, they should additionally be used to gain deeper insights into infectivity and the course of infection to develop more advanced testing strategies.

## Data Availability

All data are included in the manuscript.

## Statements

The authors declare that they have no conflicts of interest

The study did not receive any external funding.

The study was performed in agreement with the approval 122/2016 of the Ethical Committee of the Private University of Witten/Herdecke.

## References

1. Zhu N, Zhang D, Wang W, Li X, Yang B, Song J, et al. A Novel Coronavirus from Patients with Pneumonia in China, 2019. The New England journal of medicine. 2020;382(8):727–33.

2. Fineberg HV. Ten Weeks to Crush the Curve. The New England journal of medicine. 2020;382(17):e37.

3. Smithgall MC, Dowlatshahi M, Spitalnik SL, Hod EA, Rai AJ. Types of Assays for SARS-CoV-2 Testing: A Review. Laboratory medicine. 2020;51(5):e59–e65.

4. Wyllie AL, Fournier J, Casanovas-Massana A, Campbell M, Tokuyama M, Vijayakumar P, et al. Saliva or Nasopharyngeal Swab Specimens for Detection of SARS-CoV-2. The New England journal of medicine. 2020.

5. Malecki M, Lusebrink J, Teves S, Wendel AF. Pharynx gargle samples are suitable for SARS-CoV-2 diagnostic use and save personal protective equipment and swabs. Infection control and hospital epidemiology. 2020:1–2.

